# Temporal relationships between depression, self-efficacy, and physical activity in individuals with stroke

**DOI:** 10.1101/2025.04.13.25325740

**Authors:** Grace C. Bellinger, Ryan T. Roemmich, Kevin J. Psoter, Stephen T. Wegener, Eva Keatley, Margaret A. French

## Abstract

**Objective:** To investigate the temporal relationships between depressive symptoms, physical activity, and self-efficacy in individuals with stroke

**Design:** Six-month prospective observational cohort study

**Setting:** General community

**Participants:** Seventy-three individuals with stroke (42 male; 61.9±12.3 years old)

**Interventions:** Not applicable

**Main Outcome Measures:** Three functional domains were the primary outcomes: physical activity was defined by average steps per day as measured by a Fitbit device; depressive symptoms and self-efficacy were measured by Patient-Reported Outcomes Measurement Information System (PROMIS) short forms. These outcome measures were collected at study enrollment and monthly thereafter for six months, resulting in a maximum of seven timepoints.

**Results:** Three separate lagged linear mixed effects models were constructed (one with each functional domain as the dependent variable). Each model included the measure of the targeted functional domain as the dependent variable, measurements from the previous month of the two other functional domains and their interaction as fixed effects, participant as a random effect, and demographics and stroke characteristics as covariates. The depressive symptoms by self-efficacy interaction was associated with future physical activity, suggesting that higher self-efficacy positively impacts the following month’s physical activity only when depressive symptoms are low. Depressive symptoms were not associated with self-efficacy, steps per day, or their interaction in the prior month, indicating that the relationship between depressive symptoms and physical activity is unidirectional. Finally, depressive symptoms were associated with self-efficacy in the subsequent month.

**Conclusions:** The longitudinal study provides evidence that 1) mitigating depressive symptoms and promoting self-efficacy may improve future physical activity; 2) addressing depressive symptoms first may lead to more effective treatment of depression, low self-efficacy, and low physical activity; and 3) treating depression may improve future self-efficacy. Together the results provide additional knowledge about the complex relationships between mobility, mood, and self-efficacy that must be carefully managed during post-stroke rehabilitation.

## Introduction

Each year, nearly 800,000 people in the United States experience a stroke.^1^ After stroke, many individuals experience a variety of impairments related to mobility, mood, and self-efficacy. Each of these impairments are important as they impact recovery, health, and quality of life. For example, mobility limitations often lead to lower levels of physical activity, such that individuals with stroke are less active and more sedentary than adults without neurological conditions.^2–5^ This is particularly important after stroke, as physical activity can help prevent recurrent strokes and other adverse events, improve control of risk factors (e.g., diabetes, hypertension, dyslipidemia), and enhance quality of life.^6^ Similarly, post-stroke depression, which occurs in nearly one-third of stroke survivors,^7–13^ contributes to lower quality of life and poor health behaviors.^14^ Finally, self-efficacy—an individual’s belief in their ability to achieve goals, perform tasks, and/or cope with challenges^15^—is a critical factor in the rehabilitation process and commonly affected after stroke.^16,17^ Low self-efficacy can also negatively impact recovery and quality of life.^18^ To prevent negative health outcomes and improve quality of life among individuals with stroke, it is critical that mobility, depression, and self-efficacy be managed in parallel, as their impact can compound.

Although there are specific fields within rehabilitation that specialize in each of these domains (e.g., physical therapy focusing on mobility and physical activity, psychology treating depression), impairments in these domains rarely occur in isolation and are instead interrelated. As examples, prior literature in post-stroke persons has demonstrated that lower physical activity is associated with depression,^19,20^ self-efficacy and depression are inversely correlated,^21–27^ and self-efficacy relates to higher physical activity levels.^20,24,28–30^ Beyond bivariate associations, there are also important interactions to consider, as prior work has shown that self-efficacy and depression influence the relationship between physical activity and participation in individuals with stroke.^31^ Ideally, treatment plans should account for these complex relationships and guide prioritization of services among rehabilitation professionals.

The bulk of the aforementioned work exploring the complex relationships between physical activity, depression, and self-efficacy has used cross-sectional designs. While informative, cross-sectional studies do not provide evidence about the temporal, and potentially causal, relationships among these domains. Here, we aimed to characterize the temporal relationships to provide insight into causation among these three functional domains as a first step toward the larger goal of using these findings to prioritize targeted treatments of these impairments (i.e., low physical activity, low self-efficacy, or symptoms of depression). Thus, the purpose of this study was to investigate the temporal relationships between physical activity, depressive symptoms, and self-efficacy in individuals with stroke. We hypothesized that 1) depressive symptoms, self-efficacy, and their interaction would be associated with physical activity the following month and 2) physical activity would be associated with subsequent depressive symptoms.

## Methods

### Participants

The cohort used for this analysis was a convenience sample of a larger study powered for additional analyses.^32^ Participants were recruited from rehabilitation practices at Johns Hopkins Hospital between March 2021 and November 2022. Individuals were eligible for the study if they were 18 years of age or older and had a diagnosis of stroke, regardless of chronicity, as verified by chart review. Additional inclusion criteria included home wi-fi access and walking as a primary mode of mobility, though assistive devices were allowed. Given the entirely remote nature of this study, all individuals provided oral informed consent as authorized by the Institutional Review Board at the Johns Hopkins University School of Medicine (IRB00247292).

### Procedures

Participants enrolled in the prospective study for a total of six months. They were provided with a Fitbit Inspire 2 device (Fitbit Inc., San Francisco, CA, USA) and instructed to wear it continuously throughout the six-month period, only taking it off to bathe or charge the device’s battery. A custom application automatically extracted Fitbit data and, as previously described,^33^ sent automated reminders to participants instructing them to synchronize their device if it had not been synchronized for five days. From the Fitbit device, average steps per day were calculated for the 10-day window surrounding each online assessment of depressive symptoms and self-efficacy. For a window of Fitbit data to be included in the analyses, at least five of the ten days needed to be considered a valid day; a valid day was defined as any day when the device was worn for at least 75% of daytime hours (i.e., 8:00am to 9:59pm) based on the presence of heart rate data.^32^ Depressive symptoms and self-efficacy related to completing daily tasks (referred to as self-efficacy) were measured monthly with remotely administered versions of the short forms of Patient-Reported Outcomes Measurement Information System (PROMIS) Emotional Distress – Depression^34,35^ and Self-Efficacy for Managing Chronic Conditions – Manage Daily Activities,^36,37^ respectively. The PROMIS short form surveys each included four questions, which were answered using a 5-point Likert scale representing frequency of depressive symptoms or confidence in performing activities of daily living. As is conventional, the PROMIS scores for both depression and self-efficacy are reported as T-scores relative to the general population with an average of 50 and standard deviation of 10. These questionnaires were completed at enrollment and monthly thereafter, for a total of seven monthly measurements. When participants were due to complete the questionnaires, the custom application sent an email or text message, depending on participant preference, that included a participant- and time-specific link which directed participants to the testmybrain.org platform,^38,39^ where they completed the questionnaires. Participants were asked to complete the questionnaires as soon as possible but were allowed seven days to complete them.

### Statistical Analysis

Descriptive statistics of physical activity, depressive symptoms, and self-efficacy were calculated as median values and interquartile ranges. The temporal relationships between physical activity, depressive symptoms, and self-efficacy were explored using three separate lagged linear mixed effects models with participant as a random effect. The dependent variable was physical activity in Model 1, depressive symptoms in Model 2, and self-efficacy in Model 3. The independent variable lag was one month, such that in Model 1 baseline depressive symptoms and self-efficacy were paired with physical activity at month one, depressive symptoms and self-efficacy at month one were paired with physical activity at month two, and so on through month six. Similarly, in Model 2 baseline physical activity and self-efficacy were paired with depressive symptoms at month 1, physical activity and self-efficacy at month 1 were paired with depressive symptoms at month 2, and so forth. Each model included the two domains that were not the dependent variable and their interaction as fixed effects. All models were adjusted for several covariates identified a priori, including age at enrollment, sex, race, education, time since stroke in months, assistive device use, and study timepoint. All analyses were conducted in STATA Version 17.0 (StataCorp, College Station, TX, USA) using an alpha value of 0.05.

## Results

### Participants

A total of 108 adults with stroke were screened and 100 individuals were enrolled in the prospective study (Figure 1). Of those that enrolled in the study, three participants opted out of the PROMIS surveys and two participants declined the Fitbit portion; another 15 participants withdrew from the study. This resulted in 80 individuals completing the study. Since we conducted lagged analyses, the timing of the data for each functional domain was important and resulted in different sample sizes for the three models. From the 80 participants that completed the 6-month study, seven participants did not have at least one complete set of monthly measurements for any of the three models (e.g., PROMIS survey responses for one month and wearable data for the subsequent month). An additional 16 individuals did not have the required pairing for the models predicting depressive symptoms and self-efficacy. This resulted in 73 individuals [61.9±12.3 years of age, 31/73 (42.5%) female] being included in Model 1 predicting physical activity (green box in Figure 1) and 57 individuals [61.1±13.5 years of age with 23/57 (40.4%) female] being included in Models 2 and 3 predicting depressive symptoms and self-efficacy, respectively (orange box in Figure 1). Additional demographic information of the participants included in each model is shown in Table 1.

**Figure 1:**
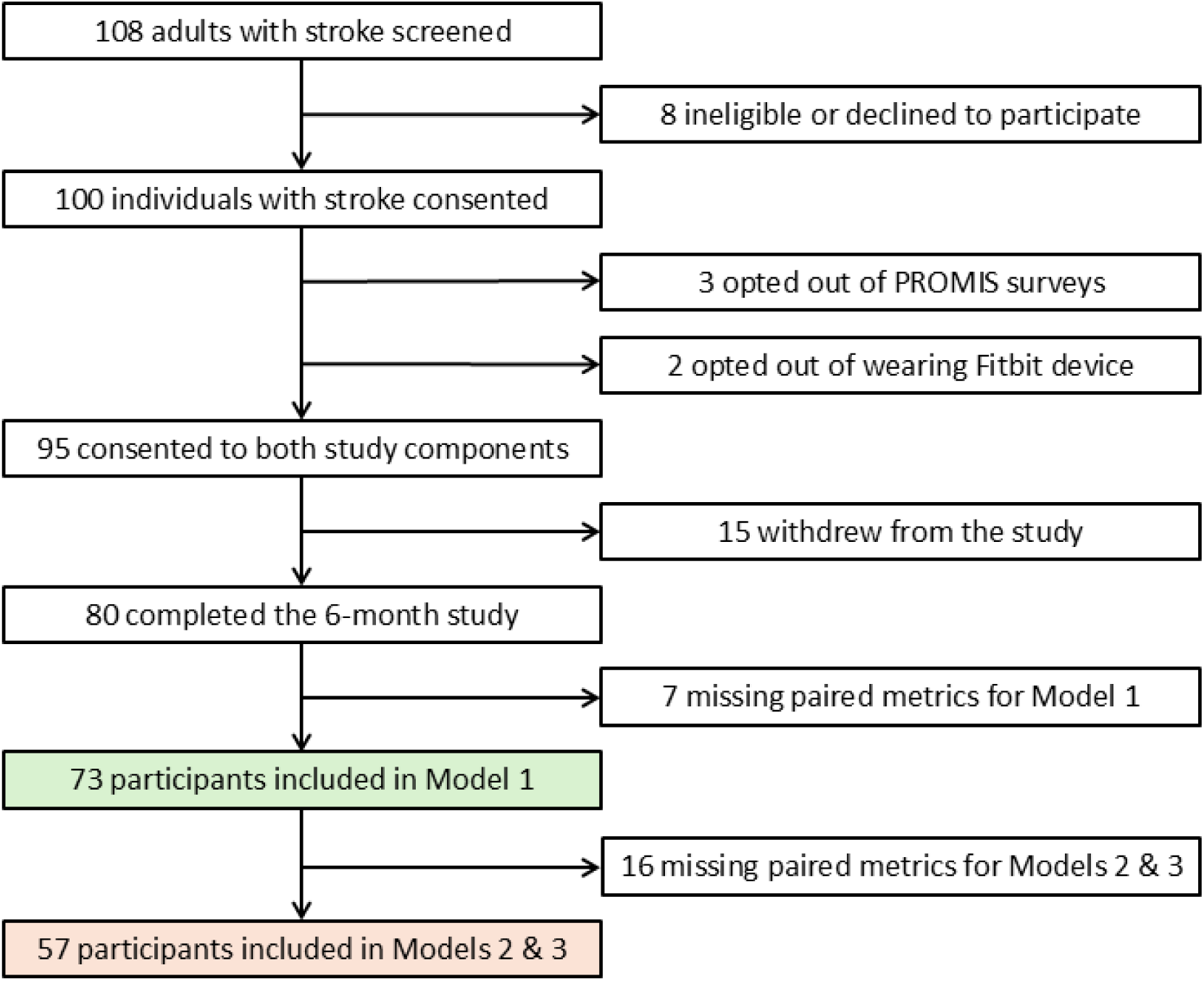
CONSORT diagram of study participants. The green box highlights the participants included in the model predicting physical activity, whereas the orange box highlights the participants included in the models predicting depressive symptoms and self-efficacy.

**Table 1:** Participant demographics and clinical characteristics for each analysis.

|  | **Model 1**<br>**(Physical Activity)**<br>**N = 73** | **Model 2 & Model 3**<br>**(Depressive Symptoms**<br>**& Self-Efficacy)**<br>**N = 57** |
| --- | --- | --- |
| **Characteristic** | **N (%)** | **N (%)** |
| Age, mean (SD) | 61.9 (12.3) | 61.1 (13.5) |
| Sex |  |  |
| Male | 42 (57.5) | 34 (59.6) |
| Female | 31 (42.5) | 23 (40.4) |
| Race |  |  |
| White | 42 (57.5) | 34 (59.6) |
| Black | 23 (31.5) | 17 (29.8) |
| Other | 8 (11.0) | 6 (10.5) |
| Education |  |  |
| College or greater | 40 (54.8) | 30 (52.6) |
| Some college | 25 (34.2) | 21 (36.8) |
| High school or less | 8 (11.0) | 6 (10.5) |
| Months since stroke at baseline, mean (SD) | 29.8 (51.6) | 31.7 (51.2) |
| <3 months | 19 (26.0) | 14 (24.6) |
| 3-6 months | 15 (20.5) | 12 (21.1) |
| 6+ months | 39 (53.4) | 31 (54.4) |
| Paretic side: Left | 41 (56.2) | 33 (57.9) |
| Assistive device use: Yes | 32 (43.8) | 27 (47.4) |

### Cohort Descriptive Statistics

The descriptive statistics summarize all available longitudinal measurements from the 73 participants included in any of the analyses. Across the 6-month study, the overall group had a median (interquartile range) physical activity of 3,995 (1,758-6,378) steps per day. The cohort’s median PROMIS depression T-score was 51.8 (41.0-57.3), with higher scores indicating more depressive symptoms. The median PROMIS self-efficacy T-score was 46.0 (40.0-59.3), where a higher score indicates greater self-efficacy.

### Physical Activity (Model 1)

Model 1 included physical activity as the dependent variable and both self-efficacy and depressive symptoms, as well as their interaction, as independent variables. After adjusting for covariates, there were significant main effects of depressive symptoms (β=201.61, p=0.02) and self-efficacy (β=298.43, p<0.01) as well as a significant interaction effect between the two (β=-4.67, p=0.01; Table 2). As shown in Figure 2, this interaction indicates that the impact of self-efficacy on subsequent physical activity differs based on depressive symptoms. Specifically, individuals with higher self-efficacy show higher levels of future physical activity when depressive symptoms are low but not if depressive symptoms are high (Figure 2). For example, at high depressive symptoms, such as the 95^th^ percentile, the predicted steps per day during the following month remain similar regardless of the self-efficacy T-score. In contrast, for individuals with low depressive symptoms, such as the 5^th^ and 25^th^ percentiles, predicted steps per day during the following month increases as self-efficacy increases.

**Figure 2:**
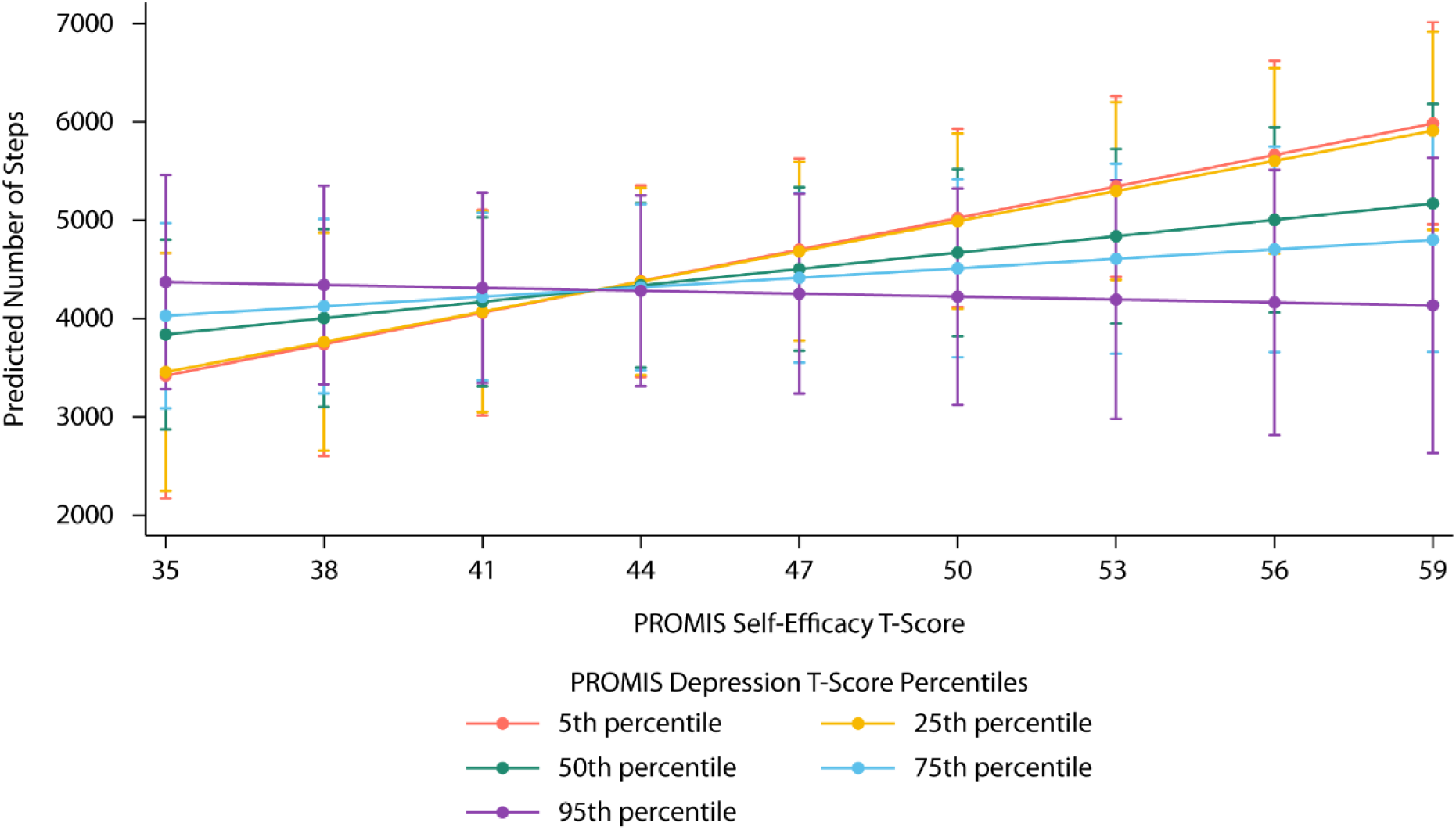
Predicted number of steps per day as a function of PROMIS self-efficacy T-scores and PROMIS depression T-score percentiles.

**Table 2:** Output of three lagged linear mixed effects models.

| Model and Variable* | Adjusted Coefficient<br>(95% Confidence Interval) | p value |
| --- | --- | --- |
| *Model 1: Physical Activity* |  |  |
| Depressive Symptoms | **201.61** steps (37.13, 366.10) | **0.02** |
| Self-Efficacy | **298.43** steps (116.53, 480.32) | **<0.01** |
| Depressive Symptoms X Self-Efficacy Interaction | **-4.67** steps (-8.15, -1.19) | **0.01** |
| *Model 2: Depressive Symptoms* |  |  |
| Self-Efficacy | -0.17 (-0.40, 0.05) | 0.13 |
| Physical Activity | 0.00 (-0.00, 0.00) | 0.62 |
| Self-Efficacy X Physical Activity Interaction | -0.00 (-0.00, 0.00) | 0.66 |
| *Model 3: Self-Efficacy* |  |  |
| Depressive Symptoms | **-0.26** (-0.41, -0.10) | **<0.01** |
| Physical Activity | 0.00 (-0.00, 0.00) | 0.68 |
| Depressive Symptoms X Physical Activity Interaction | 0.00 (-0.00, 0.00) | 0.69 |

Statistically significant adjusted coefficients and their associated p values are indicated in bold.
\*Controlling for age at enrollment, sex, race, education, time since stroke in months, assistive device use, and study timepoint

### Depressive Symptoms (Model 2)

Depressive symptoms were not significantly associated with self-efficacy (β=-0.17, p=0.13), steps per day (β=0.00, p=0.62), or the interaction between the two in the prior month (β=-0.00, p=0.66; Table 2). The absence of any significant main effects or interaction effect is illustrated in Figure 3. The lack of an offset demonstrates that there is not a significant relationship between self-efficacy and future symptoms of depression, as measured by PROMIS T-scores. The slopes near zero indicate the lack of a relationship between number of steps and depressive symptoms during the subsequent month, as within each self-efficacy T-score percentile the depression T-scores remain similar for all step count levels. The relatively parallel lines reflect the lack of a significant interaction between self-efficacy and physical activity, as the level of future depressive symptoms does not change differently based on the number of steps and self-efficacy T-score.

**Figure 3:**
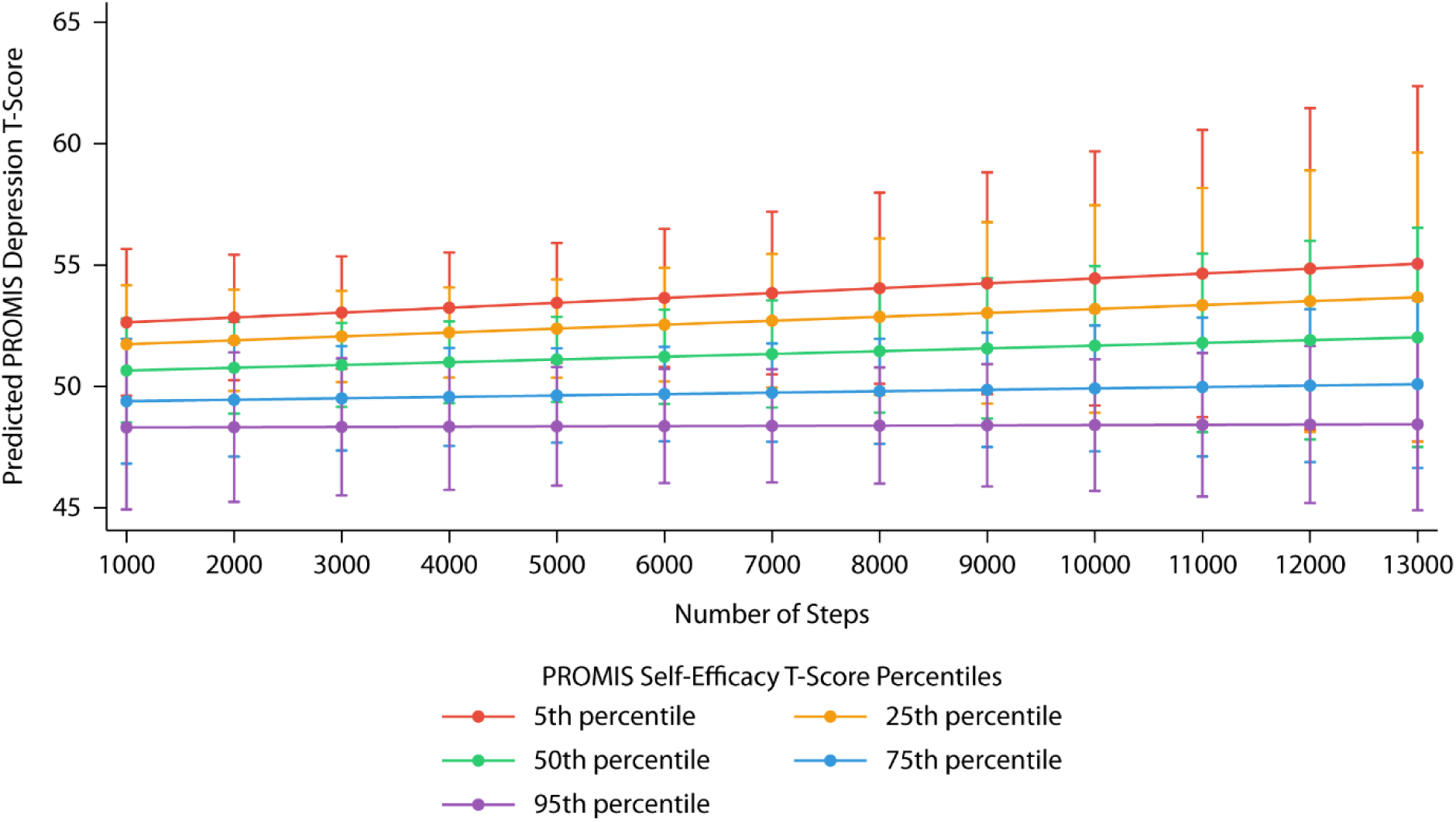
Predicted PROMIS depression T-score as a function of number of steps per day and PROMIS self-efficacy T-score percentiles.

### Self-Efficacy (Model 3)

Symptoms of depression were significantly associated with the following month’s self-efficacy (β=-0.26, p<0.01; Table 2), this is demonstrated by similarly negative slopes for all levels of physical activity in Figure 4. Physical activity, however, was not associated with future self-efficacy (β=0.00, p=0.68; Table 2). Similarly, there was not a significant interaction between depressive symptoms and physical activity (β=0.00, p=0.69; Table 2), illustrated by the relatively parallel lines. Figure 4 shows that higher depressive symptoms, regardless of physical activity level, are associated with lower self-efficacy in the subsequent month.

**Figure 4:**
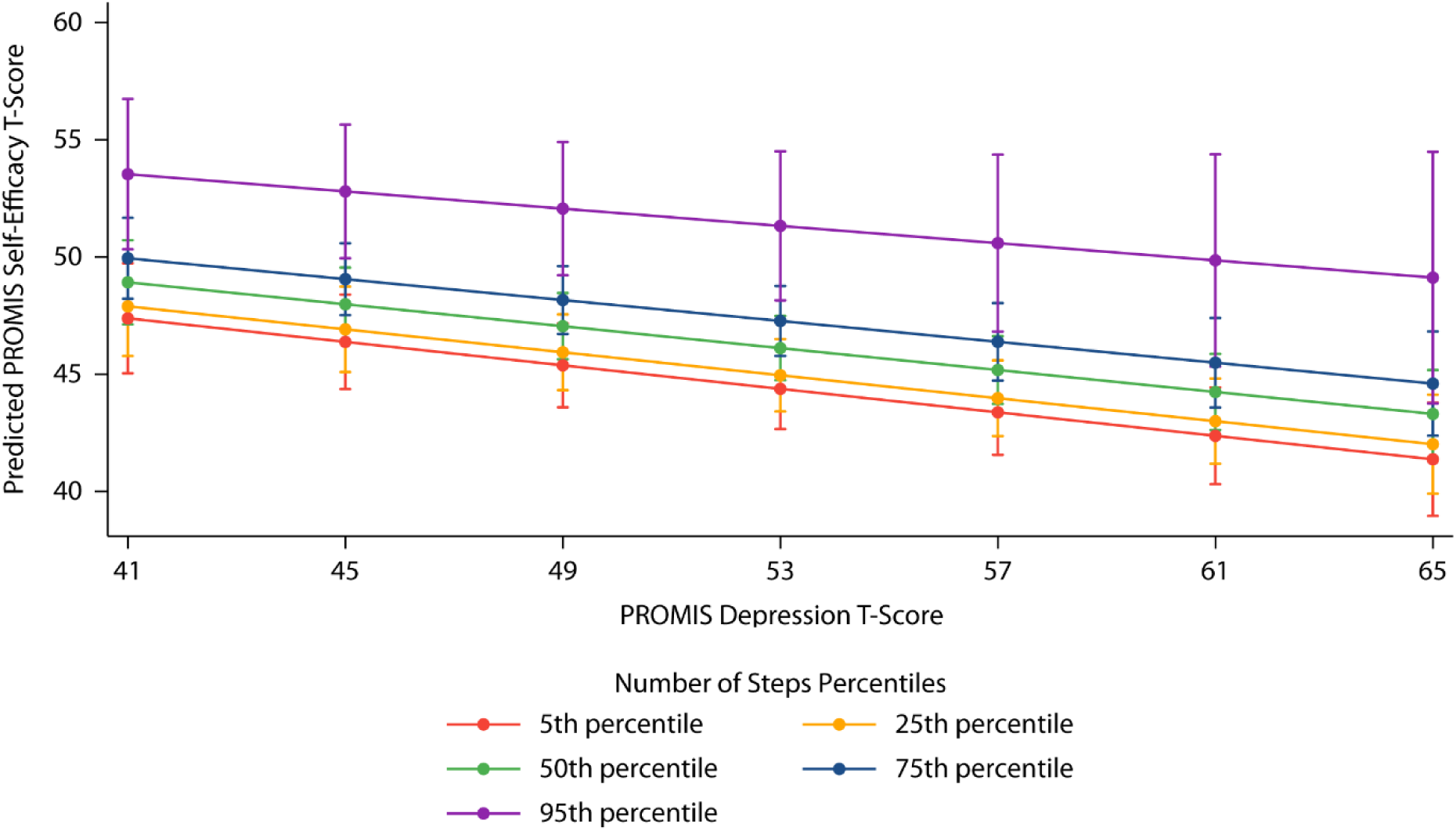
Predicted PROMIS self-efficacy T-score as a function of PROMIS depression T-scores and number of steps per day percentiles.

## Discussion

The purpose of this study was to explore temporal relationships between physical activity, depressive symptoms, and self-efficacy in individuals with stroke. The lagged linear mixed effects models provide insight into potential causal relationships between these three functional domains, as well as their interactions, since the dependent variable in these models occurred in the future. Understanding these complex relationships can inform rehabilitation professionals’ efforts to time and prioritize treatments that address these impairments across different domains. The findings suggest that, when present, treating depressive symptoms first may facilitate more effective interventions targeting low physical activity and low self-efficacy.

There was a significant interaction between self-efficacy and depressive symptoms associated with physical activity the subsequent month. Depressive symptoms and self-efficacy interact such that higher self-efficacy can boost future physical activity in individuals with low symptoms of depression. However, if depressive symptoms are high, physical activity is low regardless of self-efficacy. The significant interaction between depressive symptoms and self-efficacy being associated with subsequent physical activity is consistent with cross-sectional literature demonstrating that lower levels of depression and higher levels of self-efficacy are associated with higher steps per day and increased overall physical activity in individuals with stroke.^19,20,28,31^ Additionally, previous work had shown that depression in acute stroke correlates with reduced physical activity in the future; however, they did not account for self-efficacy.^40^ Our findings add to this understanding due to the longitudinal nature of our data and the inclusion of all three functional domains. While we did not test this in the current study, our results suggest that to increase physical activity, clinicians may aim to both treat depression and enhance self-efficacy in addition to directly addressing physical inactivity. Particularly for those with high depressive symptoms, intervening on the mood impairment first may provide more effective and holistic treatment. Future work that assesses the timing of addressing these functional domains would assist in translating our findings into clinical practice.

Neither physical activity nor self-efficacy impacted future depressive symptoms. While previous literature has demonstrated cross-sectional correlations between depression and physical activity,^19,20^ the current findings do not support a bidirectional temporal relationship. Similarly, prior work has demonstrated cross-sectional correlations between depression and self-efficacy,^22,23,25,26^ which were not observed in this study. Our results are in contrast to previous literature suggesting general self-efficacy approximately eight weeks post-stroke influences depression six months post-stroke^27^ and sedentariness in the acute hospitalization is associated with depression three months post-stroke.^41^ However, these discrepancies may be due in part to the current analyses using a one-month lag and including individuals with chronic stroke. Though depressive symptoms were significantly associated with future physical activity and self-efficacy in Models 1 and 3, the temporal relationships were not bidirectional. The absence of any significant main or interaction effects in Model 2 demonstrates that future depression may not be associated with variables accounted for in the current study.

Self-efficacy was associated with prior depressive symptoms but not past physical activity. The influence of depressive symptoms builds on previous work demonstrating associations between depression and self-efficacy^22,23,25–27^ and extends the body of evidence toward a causal relationship.^42^ The results of Models 1 and 3 suggest that mitigating depressive symptoms through targeted intervention may improve not just depression, but also future self-efficacy and physical activity. Though prior literature has shown cross-sectional relationships between physical activity and self-efficacy,^20,28^ the current work suggests that self-efficacy, through its interaction with depression, influences future physical activity, but there is no evidence for the reverse temporality.

Together, the results of the three models suggest the following: 1) mitigating depressive symptoms and promoting self-efficacy, in that order, may improve future physical activity, 2) future depressive symptoms may not be associated with physical activity or self-efficacy, and 3) treating symptoms of depression may improve future self-efficacy. The cohort of 73 individuals with stroke was representative of typical post-stroke persons in terms of objective physical activity and self-reported depressive symptoms and self-efficacy, improving the generalizability of our findings. The group of participants collectively took approximately 4,000 steps per day across the 6-month study, which is less than the amount of physical activity recommended by the Department of Health and Human Services for American adults^43^ as well as exercise recommendations specific to post-stroke persons.^6^ Despite the prevalence of post-stroke depression,^7–13^ the cohort self-reported depressive symptoms similar to the general public, as evidenced by a median T-score close to 50. In alignment with prior work,^16,17^ the individuals with stroke scored slightly lower on self-efficacy than normative values, which would be a T-score of 50.

### Study Limitations

This longitudinal study does have a few important limitations. The PROMIS depression short form survey was used to measure depressive symptoms rather than a clinical diagnosis of depression. While self-reported, the use of PROMIS surveys allowed for repeated measures on a more continuous scale than a diagnosis would provide. Another limitation is that the current analysis used a one-month lag for the lagged linear mixed effects models. The one-month lag was chosen based on the administration of monthly PROMIS surveys, though two- or three-month lags could be used to explore temporal relationships over longer timescales. Lastly, the findings provide evidence for temporal relationships but are insufficient to prove causality. Though temporality is an important component of causality, causal relationships require additional criteria not explored in this study. While the results of this study provide temporal evidence in support of causality, future work should explore the other key criteria.

## Conclusions

The findings of this longitudinal observational study have clinical implications for the multi-disciplinary holistic treatment of individuals with stroke. To promote physical activity after stroke, both depression treatment and self-efficacy enhancement may be necessary in addition to interventions addressing physical inactivity or sedentary behavior. For individuals with stroke with low physical activity, high depression, and low self-efficacy, targeting physical activity in isolation may be less effective and may not address the other two areas of concern. Addressing depressive symptoms, however, has the benefit of mitigating depression while also promoting improvement in both self-efficacy and physical activity.

## Data Availability

All data produced in the present study are available upon reasonable request to the authors.

## Acknowledgments

This work was funded by the National Institutes of Health (NCMRR F32HD108835-01 awarded to MAF), the American Heart Association (24POST1187285 awarded to GCB), and the Sheikh Khalifa Stroke Institute at Johns Hopkins Medicine. The authors thank Junyao Li for his assistance with participant recruitment and data collection.

## Notes

### Competing Interest Statement

The authors have declared no competing interest.

### Author Declarations

The Institutional Review Board at the Johns Hopkins University School of Medicine gave ethical approval for this work.

